# Pathway specific polygenic risk scores identify pathways and patient clusters associated with inflammatory bowel disease risk, severity and treatment response

**DOI:** 10.1101/2021.11.19.21266549

**Authors:** Corneliu A. Bodea, Michael Macoritto, Yingchun Liu, Wenliang Zhang, Jozsef Karman, Emily A. King, Jacob F. Degner, Marc C. Levesque, J. Wade Davis, Sherry Cao

## Abstract

Crohn’s disease (CD) and ulcerative colitis (UC) are inflammatory bowel diseases (IBD) with a strong genetic component [1,2]. Genome-wide association studies (GWAS) have successfully identified over 240 genetic loci that are statistically associated with risk of developing IBD, and these associations provide valuable insights into disease pathobiology. Building on GWAS findings, conventional polygenic risk scores (cPRS) aim to quantify the aggregated disease risk based on DNA variation, and these scores can identify individuals at high risk. While stratifying individuals based on cPRS has the potential to inform clinical care, the development of novel therapeutics requires deep insight into how aggregated genetic risk leads to disruption of specific biological pathways. Here, we developed a pathway-specific PRS (pPRS) methodology to assess IBD common variant genetic risk burden across 31 manually curated pathways. We first prioritized 206 genes based on comprehensive fine-mapping and eQTL colocalization analyses of genome-wide significant IBD GWAS loci and 58 highly penetrant genes based on their involvement in early onset IBD or autoimmunity-related colitis. These 264 genes were assigned to at least one of the 31 pathways based on Gene Ontology annotations and manual curation. Finally, we integrated these inputs into a novel pPRS model and performed an extensive investigation of IBD disease risk, severity, complications, and anti-TNF treatment response by applying our pPRS approach to three complementary datasets encompassing IBD cases and controls. Our analysis identified multiple promising pathways that can inform drug target discovery and provides a patient stratification method that offers insights into the biology of treatment response.

## Introduction

Inflammatory bowel diseases (IBD) are a group of chronic complex relapsing and remitting disorders that involve a dysregulated immune response leading to debilitating digestive tract inflammation [3]. IBD is usually subdivided into two categories: Crohn’s disease (CD) and ulcerative colitis (UC), where UC mainly affects the colon and CD can affect any region of the digestive tract. Risk of developing IBD has long been recognized as having a genetic as well as an environmental component, with up to 12% of IBD patients having a family history of IBD, and both adult-onset and early-onset forms of IBD are observed [1,2].

Early-onset IBD forms are typically driven by inborn errors of immunity, and multiple genes have been recorded with rare, highly penetrant coding variants that lead to early-onset IBD or autoimmune disease with IBD manifestations [4,5,6]. Examples include genes within the IL10 pathway such as IL10RA and IL10RB. In addition to highly penetrant genes involved in early-onset IBD or colitis-associated autoimmune disease, genome-wide association studies (GWAS) have reported 241 common genetic loci associated with IBD disease risk [7]. These loci provide a valuable insight into IBD biology and enable a prioritization of individual genes with likely involvement in disease processes.

Two major challenges have affected the direct translation of GWAS results to novel insights into disease pathogenesis and new therapeutic options. First, it is difficult to pinpoint causal disease variants from GWAS results due to linkage disequilibrium, because neighboring highly correlated variants often share similar evidence for GWAS disease association. Second, the majority of GWAS signals fall in noncoding regions of the human genome and it is thus unclear how they are mechanistically linked to disease. To address these challenges and identify genes with higher evidence for causality, statistical fine-mapping methods can be applied to resolve causal variants even in loci with high linkage disequilibrium [8]. In addition, statistical colocalization methods can connect noncoding signals with potential target genes by testing the overlap between GWAS peaks and expression quantitative trait loci (eQTL) variants that are associated with altered mRNA expression of neighboring genes [9].

While current insights into IBD biology derived from cases of early disease onset and GWAS have served as a powerful starting point for experimental elucidation of disease mechanisms, evaluating disease risk at the individual patient level has the potential to inform clinical care, to enable early detection, and to identify effective prevention methods [10]. Conventional polygenic risk scores (cPRS) aim to quantify cumulative genetic disease predisposition via a weighted sum of risk-associated alleles at common variants across the genome. A challenge with cPRS is that a unified genome-wide risk score lacks the granularity to highlight individual biological pathways contributing to disease biology in individual subjects. Multiple pathway-level PRS (pPRS) models have been proposed to address this challenge [11,12,13], however the connection of variants to target genes is still mainly limited to gene body regions or coding variants, and no uniform approach has been proposed for the selection of genes that are both representative for individual biological pathways as well as relevant for the phenotype under consideration.

Here, we present a novel pPRS methodology that aims to quantify genetic risk at the biological pathway level and enables patient stratification based on individual genetic risk profiles (Figure 1). To prioritize a list of genes with likely involvement in IBD we first performed comprehensive statistical fine-mapping and colocalization analyses on all genome-wide significant loci from the latest IBD GWAS [7] and integrated these findings with published results in other IBD cohorts or relevant cell types. We prioritized 206 genes based on our analysis of IBD GWAS loci and 58 genes due to their involvement in early onset IBD or in monogenic disease with IBD manifestations. We organized this collection of genes into 31 pathways using GO annotations and manual curation. Finally, we leveraged Open Targets variant-to-gene scores [14] to connect individual GWAS variants to these curated genes and pathways and leveraged these connections to compute pPRS scores.

**Figure 1.**
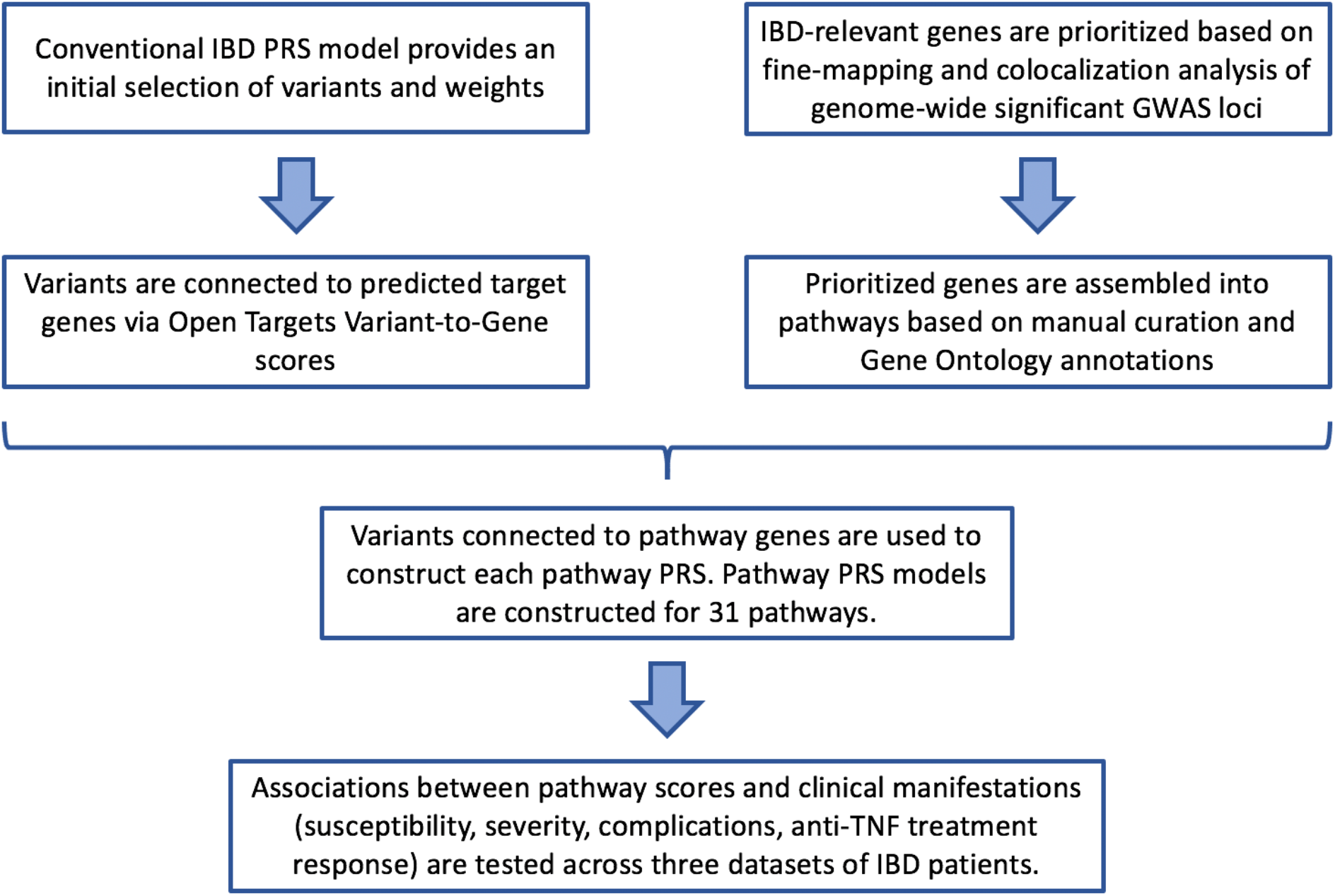
Study design and workflow. A pPRS method was developed to expand on conventional PRS models starting from a prioritized list of IBD-relevant genes. Genes were prioritized based on statistical colocalization and fine-mapping analyses applied to published IBD GWAS results, and based on involvement in early-onset IBD or colitis-associated autoimmune disease. These genes were grouped into pathways, and variants were connected to pathway genes based on genetic and epigenetic evidence curated and integrated by the Open Targets consortium. As with a conventional PRS, pPRS values were constructed by summing up genetic risk across multiple variants. However, each pPRS model only relied on variants relevant to that pathway based on each variant’s connection to pathway genes. Finally, we tested the association between our results across 31 pathways and clinical characteristics of IBD patients in three datasets.

Our goal was to evaluate our new pPRS methodology by performing an in-depth investigation of IBD biology across multiple well-powered patient datasets. We first applied our pPRS method to a collection of over 3000 IBD cases and ancestry-matched controls from the UK Biobank [15], a large-scale resource encompassing genetic and in-depth phenotypic information on approximately 500,000 UK participants. We found significant associations between individual pathways and disease status, as well as subpopulations defined by pPRS profiles that are enriched for IBD cases. We then computed pPRS values for 790 CD cases from the RISK consortium [16]. Here we identified significant correlations between pPRS-derived patient clusters and disease severity at study enrollment. Finally, we leveraged our pPRS methodology to identify significant biological mechanisms associated with IBD complications and anti-TNF treatment response in the PANTS cohort [17], a prospective observational UK-wide study of CD patients under anti-TNF treatment. Based on pPRS values as well as environmental factors we constructed PANTS patient clusters that show higher fractions of responders or non-responders compared to the study-wide proportions, suggesting the utility of the pPRS metric as a tool for patient stratification. To our knowledge this represents the most comprehensive PRS analysis of IBD to date.

## Material and Methods

### Prioritization of genes with evidence for involvement in IBD susceptibility

To identify genes with strong supporting evidence for involvement in IBD susceptibility we performed statistical fine-mapping and colocalization analyses on the latest published IBD GWAS [7]. We identified the independent signals at all 241 reported IBD GWAS loci via the GCTA pipeline (version 1.93.2beta) [18] and then performed statistical fine-mapping of all independent signals by computing Bayes factors and calculating 95% credible sets. Genes that contain coding variants with posterior probability greater than 50% were recorded for the pathway PRS analysis. This posterior probability threshold enabled identification of coding variants that capture the majority of the genetic association evidence at their locus.

Colocalization of GWAS and eQTL signals was tested via the coloc R package (R version 4.0.2, coloc version 3.2-1) [9]. We extracted eQTL data from GTEx release V8 [19] and restricted our analysis to ten IBD-relevant tissues and cell types: Small Intestine Terminal Ileum, Stomach, Whole Blood, Esophagus Mucosa, Colon Transverse, Cells EBV-transformed lymphocytes, Cells Cultured fibroblasts, Esophagus Gastroesophageal Junction, Colon Sigmoid, and Esophagus Muscularis. Genes with colocalization probability greater than 80% (based on the coloc PP4 value) were recorded for the pathway PRS analysis. The colocalization probability threshold of 80% enabled a stringent selection of genes with both GWAS and eQTL evidence and was used in previous colocalization studies [9].

We then assembled a list of genes with known clinical involvement in early onset IBD or in immune disease with IBD manifestations (for example severe diarrhea) based on published evidence [4,5,6]. The final list of IBD-relevant genes used in the construction of pathways contained 264 genes.

### Biological categorization of the IBD Very Early Onset, Early Onset and GWAS genes

Very Early Onset (VEO) and Early Onset (EO) genes lists from published studies [4,5,6] were interconnected using the protein-protein interaction tool STRING [20], allowing for one protein mediator to be added via a shortest path algorithm to enable connectivity (Figure 2A). After the generated graphs connected different groups of genes in a proximal manner, the functionality of each gene was reviewed in the literature and Gene Ontology (GO) [21,22] overrepresentation of biological function analysis was performed. Gene groups were then assigned the most prevalent biology based on the results (Figure 2B). For the high-confidence GWAS genes, all were used for GO overrepresentation of Biological Process analysis and GO terms of very similar function were grouped together based on manual curation and summarized to give a granular view of functionality. These functions were then grouped together two more times in sequence to give a medium and high-level grouping of the biology. At each level, the GWAS genes associated with the biological categorizations were tracked.

**Figure 2.**
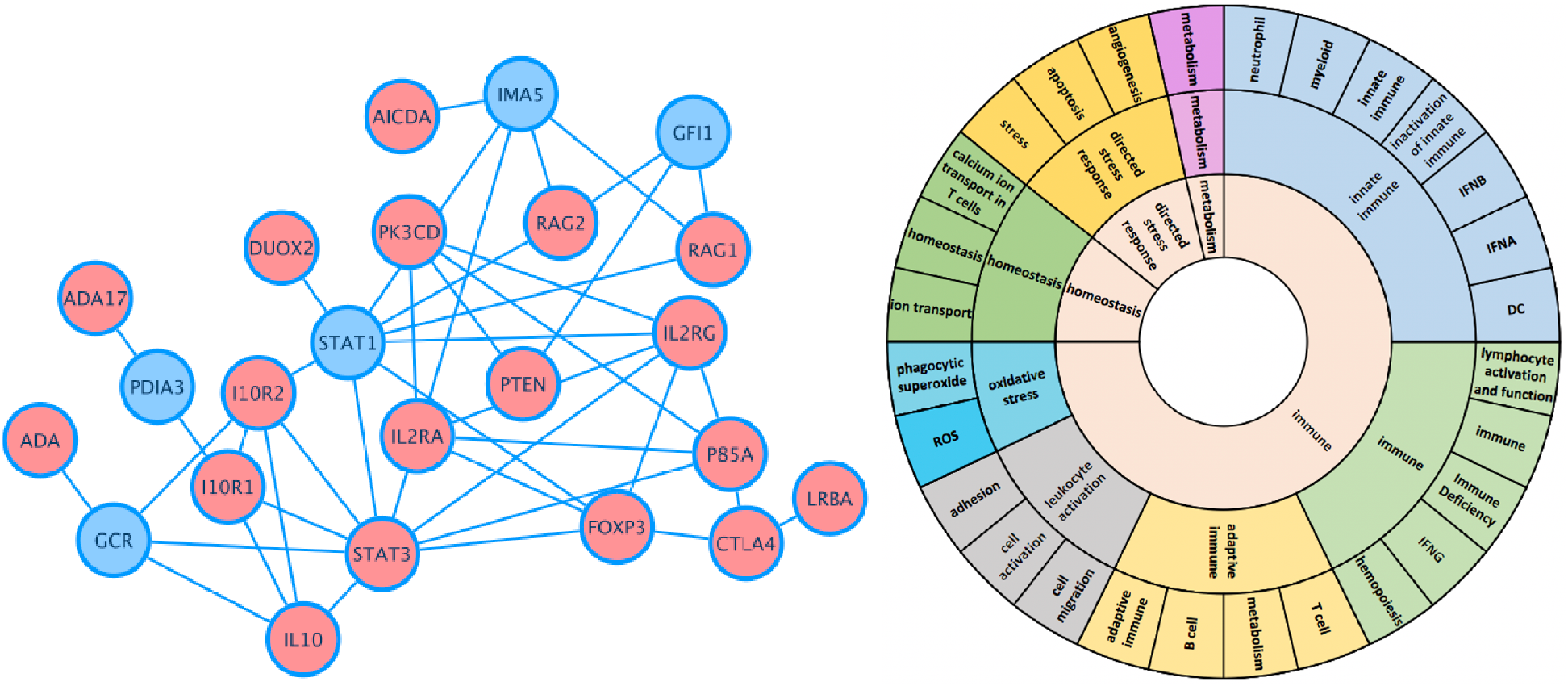
Biological pathways represented in the set of 264 IBD-implicated genes. (A) Subnetwork showcasing IBD-implicated genes (red) and protein mediators (blue) (B) We grouped genes at three levels of increasing granularity, with genes being allowed to reside in multiple pathways. For the remainder of our work, we focused on the most granular pathway assignments when constructing pPRS metrics (outermost ring).

### Construction of the pPRS model for IBD

We extracted the variants and weights from an IBD cPRS model which was trained on 120,280 individuals that were part of the UK Biobank phase 1 dataset [10]. To connect variants to their biologically relevant genes we leveraged the variant-to-gene (V2G) scores developed by Open Targets (latest version downloaded on March 4^th^, 2020) [14]. These scores range from 0 (weak) to 1 (strong) and quantify the strength of evidence connecting a variant to a particular gene based on QTL data, chromatin conformation experiments, and in silico functional prediction. The genetic burden contributed by a variant to a pathway gene is computed as the product of allele dosage, cPRS coefficient (based on GWAS effect size), and confidence that the variant affects gene function (Figure 3). The pPRS score is then constructed by summing up the individual variant-level products across all variants that map to genes in the pathway of interest (based on non-zero V2G scores):

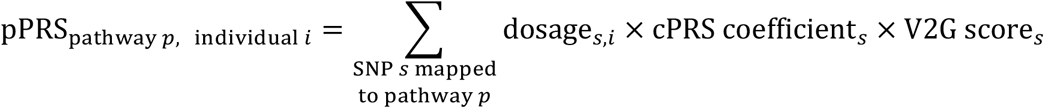

**Figure 3.**
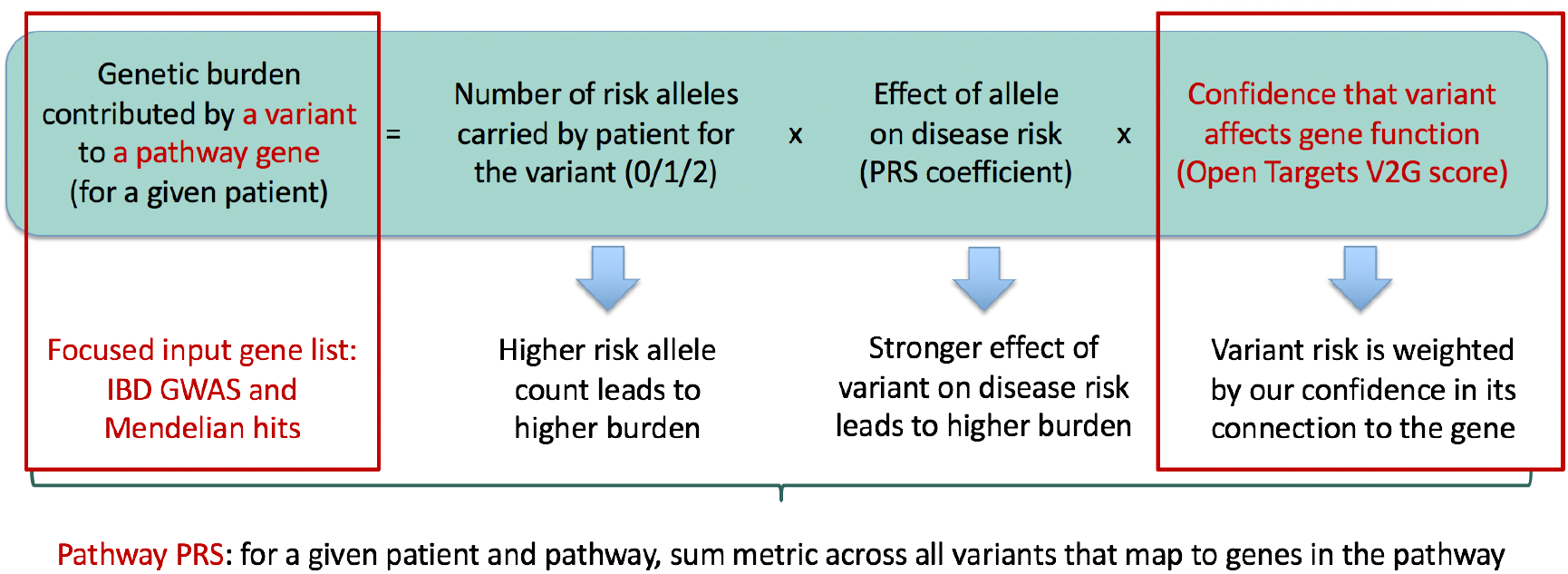
Construction of pPRS scores. Variants are connected to genes via the continuous Open Targets V2G score (range between 0 and 1) that integrates transcriptomic, epigenetic as well as functional variant-level evidence. As in a cPRS, dosage is multiplied by the corresponding GWAS-derived variant weight, and this is further scaled by the V2G score to estimate the genetic burden contributed by a variant to a pathway gene. Finally, these values are added across all variants connected to genes in a pathway (based on a nonzero V2G score) to construct the pPRS metric for a given sample. This process is repeated for all pathways identified in our IBD-focused input gene set. Steps of our scoring procedure marked with red boxes represent novel elements emphasized in our pPRS approach.

This approach has the advantage of leveraging a continuous variant-to-gene confidence metric across all relevant variants in the genome (coding as well as noncoding). Starting from an IBD-centered input gene list also helps reduce potential noise by prioritizing only genes with the strongest evidence for involvement in IBD biology.

### Cohorts with clinical and genetic data

We computed cPRS and pPRS values in three cohorts that offer complementary insights into IBD biology. We first leveraged IBD cases and controls from the UK Biobank [15] to evaluate the performance of the pPRS model in a disease susceptibility dataset. The UK Biobank is a large biomedical database that contains in-depth health information as well as genotyping data for approximately 500,000 UK participants. Genotyping QC, imputation and ancestry mapping were performed as part of the UK Biobank study. Next, we studied a cohort of early-onset IBD cases from the RISK study [16] to evaluate the association between pathway PRS values and disease severity. RISK is a 36-month prospective inception cohort study that has collected data from over 1000 pediatric Crohn’s disease patients from 28 clinics across the United States and Canada. Finally, IBD patient data from the personalized anti-TNF therapy in Crohn’s disease study (PANTS) [17] was leveraged to connect pPRS values to IBD complications as well as anti-TNF treatment response. PANTS is a prospective observational UK-wide study that enrolled anti-TNF naïve Crohn’s disease patients. After anti-TNF treatment, patients were evaluated for 12 months or until drug withdrawal, and clinical as well as genetic data was collected at baseline.

### Testing the association between pathway PRS values and disease traits

Three types of statistical analyses were performed to connect pPRS values to clinical observations. We first applied two-sided t-tests to check for differences in the pPRS distributions in IBD cases versus controls. Second, we leveraged elastic net regression to model all pathways together while accounting for the correlation structure caused by shared genes. These regularized models highlight individual pathways by identifying the strongest associations with our clinical phenotypes (susceptibility, severity, treatment response). The elastic net loss function has two parameters:

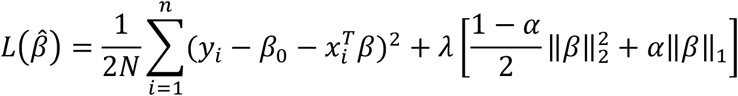

The parameter *λ* controls the overall level of regularization while the parameter *α* controls the mixing between ridge (*α* = 0) and lasso regression (*α* = 1). We tuned both parameters via a grid search and 10-fold cross-validation as implemented in the R caret package (R version 4.0.2, caret version 6.0-86). All features were also scaled and centered before running the elastic net regression to ensure effective regularization. Finally, we performed hierarchical clustering on our sample cohorts based on pPRS values. To test for enrichment of clinical characteristics (for example IBD cases) within individual sample clusters we used a hypergeometric model: enrichment p-value = *P*(*X* ≥ *x*), where 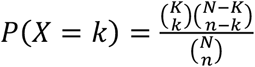 represents the hypergeometric probability mass function.

## Results

### Pathway analysis of IBD susceptibility

To evaluate whether the pPRS approach can identify statistically significant differences between IBD cases and controls, we leveraged samples from the UK Biobank [15]. The UK Biobank is a large biomedical database that contains in-depth health information as well as genotyping data for 500,000 UK participants. Genotyping QC, imputation and ancestry mapping were performed as part of the UK Biobank study. Based on the available health records we identified 8,190 IBD cases (white British ancestry, K50 or K51 ICD10 codes). We also identified 215,677 ancestry-matched individuals of age 55 or higher with no recorded autoimmune or fibrotic disease (including IBD). After excluding UK Biobank phase 1 participants (which were used in the training of the original cPRS model [10]) as well as individuals with missing genotyping data we included 3,278 IBD cases and 3,373 controls for the PRS analysis.

We first computed cPRS values for all selected samples and compared the scores across IBD subcategories (Figure 4).

**Figure 4.**
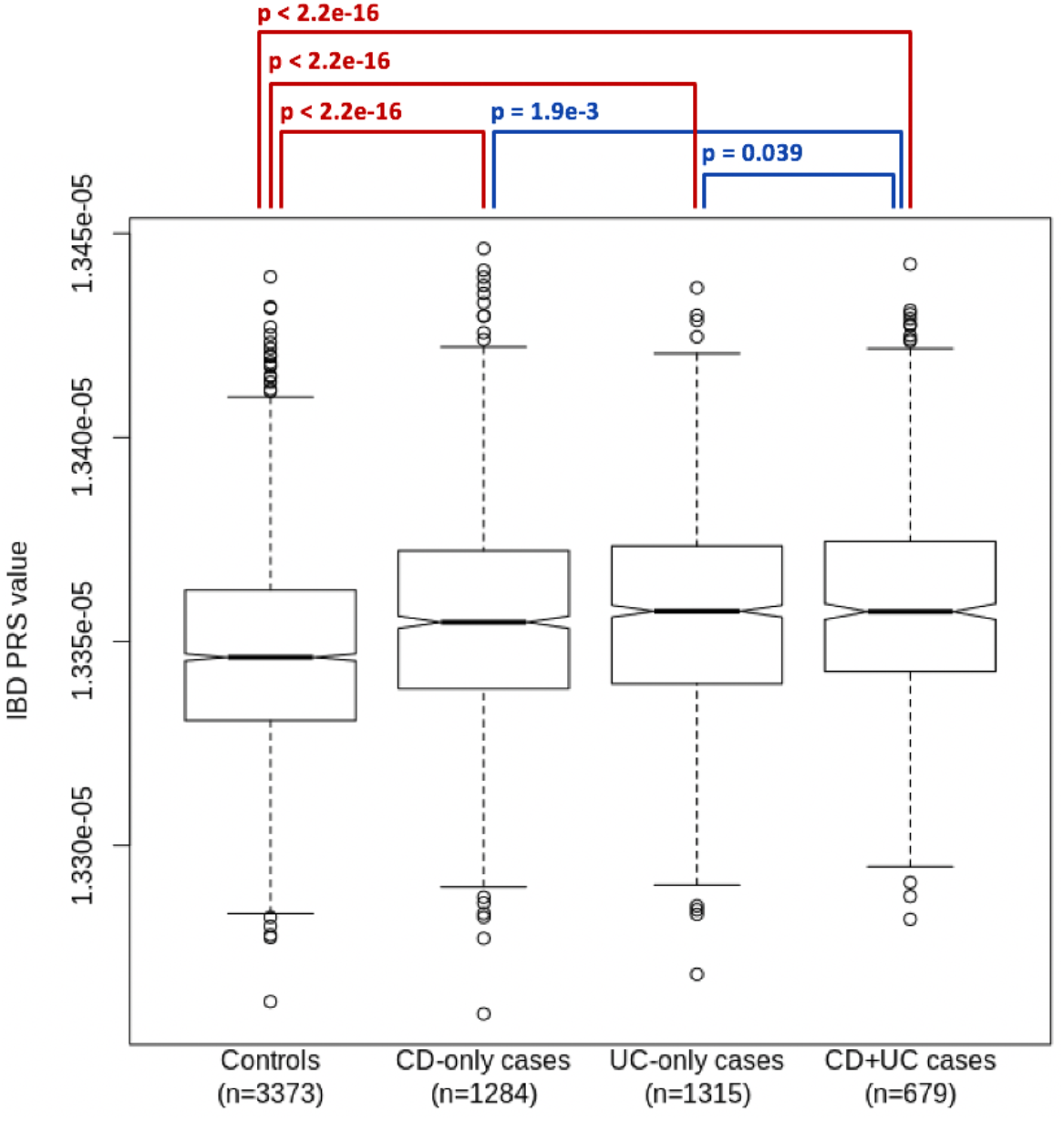
Conventional IBD PRS scores for UK Biobank IBD patients and controls. Significantly lower scores are observed in controls versus cases. Lower cPRS values are also present in CD-only or UC-only cases versus UK Biobank participants with both CD and UC diagnoses. Significance levels are based on one-sided t-tests.

As expected, controls had lower cPRS values than IBD patients (one-sided t-test p-value < 2.2e-16). Patients with CD-only or UC-only diagnoses also had lower cPRS values than patients diagnosed with both CD and UC. This confirms the utility of cPRS to identify subtle differences in genetic susceptibility between disease and healthy contexts. While such subtle differences limit the overall predictive ability of cPRS, subsets of samples enriched for cases and controls can be identified based on the extremes of the cPRS distribution. For example, the bottom 20% (n = 1329) of cPRS values included 36% IBD cases, whereas the top 20% (n = 1328) of the cPRS values included 66% IBD cases.

To compare the pPRS values between IBD cases and population controls we performed two-sided t-tests across all 31 pathways (Table 1). Most pathways (25 out of 31) exhibited significantly higher scores in the IBD case cohort, even after Bonferroni multiple testing correction. This result was expected based on the cPRS signal and the selection of pathway genes, and it confirms that pPRS can detect biologically relevant differences between disease states and recover the expected directionality of the genetic burden (higher burden in IBD cases compared to controls).

**Table 1.**
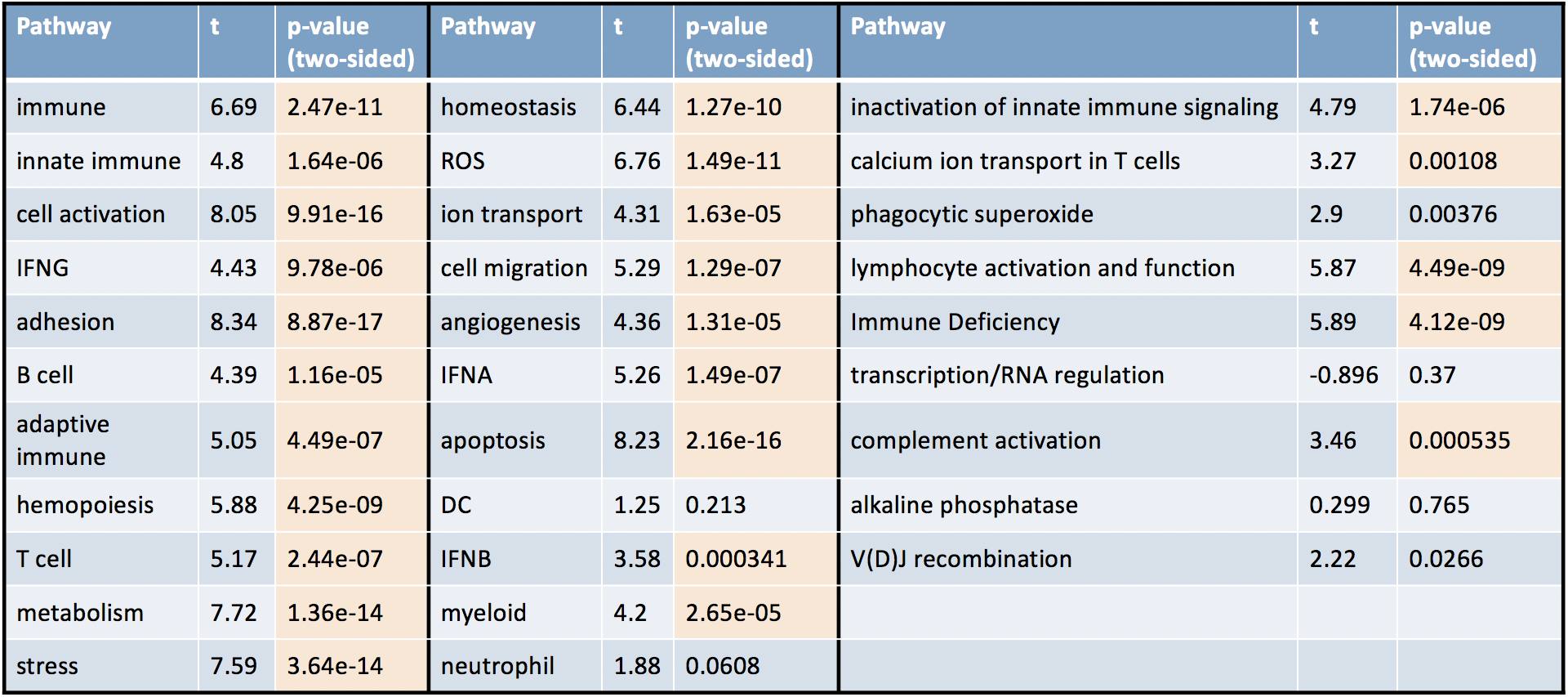
Differences in pathway PRS distributions between UK Biobank IBD cases and controls. Results from two-sided t-tests are displayed, together with the corresponding t-statistics. Orange entries represent p-values significant after Bonferroni correction for 31 pathway tests. Most pathways (25 out of 31) have significantly higher genetic risk burden in cases compared to controls, which falls in line with biological expectations and confirms the utility of pPRS in a case versus control comparison.

| Pathway | t | p-value<br>(two-sided) | Pathway | t | p-value<br>(two-sided) | Pathway | t | p-value<br>(two-sided) |
| --- | --- | --- | --- | --- | --- | --- | --- | --- |
| immune | 6.69 | 2.47e-11 | homeostasis | 6.44 | 1.27e-10 | inactivation of innate immune signaling | 4.79 | 1.74e-06 |
| innate immune | 4.8 | 1.64e-06 | ROS | 6.76 | 1.49e-11 | calcium ion transport in T cells | 3.27 | 0.00108 |
| cell activation | 8.05 | 9.91e-16 | ion transport | 4.31 | 1.63e-05 | phagocytic superoxide | 2.9 | 0.00376 |
| IFNG | 4.43 | 9.78e-06 | cell migration | 5.29 | 1.29e-07 | lymphocyte activation and function | 5.87 | 4.49e-09 |
| adhesion | 8.34 | 8.87e-17 | angiogenesis | 4.36 | 1.31e-05 | Immune Deficiency | 5.89 | 4.12e-09 |
| B cell | 4.39 | 1.16e-05 | IFNA | 5.26 | 1.49e-07 | transcription/RNA regulation | -0.896 | 0.37 |
| adaptive immune | 5.05 | 4.49e-07 | apoptosis | 8.23 | 2.16e-16 | complement activation | 3.46 | 0.000535 |
| hemopoiesis | 5.88 | 4.25e-09 | DC | 1.25 | 0.213 | alkaline phosphatase | 0.299 | 0.765 |
| T cell | 5.17 | 2.44e-07 | IFNB | 3.58 | 0.000341 | V(D)J recombination | 2.22 | 0.0266 |
| metabolism | 7.72 | 1.36e-14 | myeloid | 4.2 | 2.65e-05 |  |  |  |
| stress | 7.59 | 3.64e-14 | neutrophil | 1.88 | 0.0608 |  |  |  |

Due to overlapping genes between pathways we observe a strong correlation structure in pPRS scores (Figure S1). Given these correlation patterns we investigated which pathways provide the most power to differentiate between IBD cases and controls. We performed elastic net logistic regression (Material and methods) and identified 8 prioritized pathways (Figure 5). Consistent with the individual pathway t-test results, all regression coefficients were positive, indicating that higher genetic risk burden over these pathways correlated with higher disease susceptibility.

**Figure 5.**
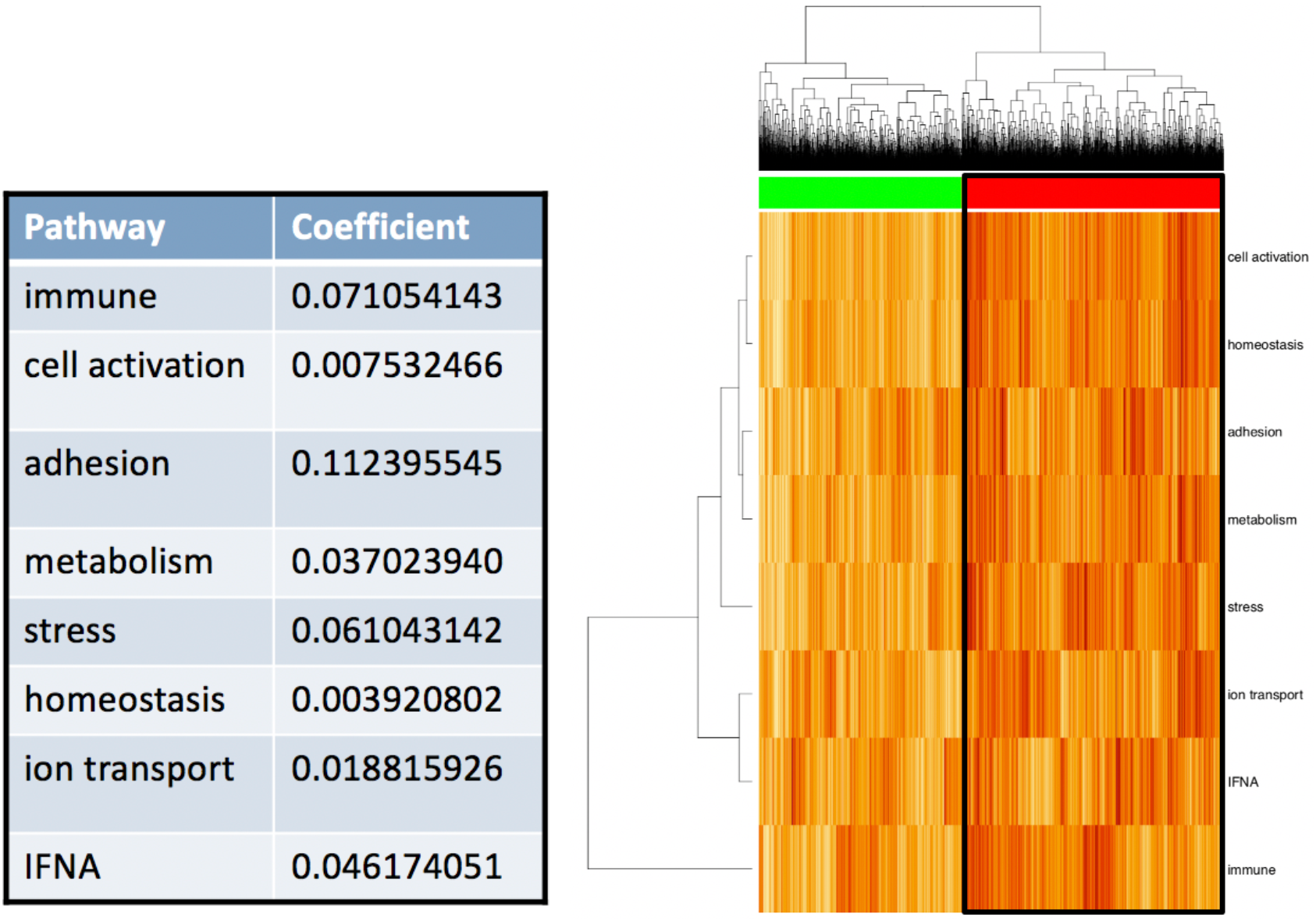
Most predictive IBD susceptibility pathways identified by elastic net logistic regression. Eight pathways were prioritized by the regularized regression model, with all pathways showing a positive association between genetic risk and IBD susceptibility. Hierarchical clustering based on these eight pathways identified a UK Biobank cohort enriched for IBD cases (red cluster: hypergeometric test p-value = 6.9e-13).

Starting from these prioritized pathways we next evaluated whether the pPRS metric could be used to construct sample subsets enriched for IBD cases (Figure 5). We performed hierarchical clustering on the UK Biobank cases and controls based on their pPRS values and identified a significant over-representation of IBD cases (hypergeometric test p-value = 6.9e-13) after the first level of the clustering hierarchy. This observation was consistent with the elastic net results and served as a confirmation that sample clusters with statistically significant phenotypic enrichments could be constructed based on the pPRS methodology.

### Pathway analysis of IBD severity

To expand our use of pPRS metrics beyond disease susceptibility we next applied our methodology to a cohort of early-onset IBD cases from the RISK study [16]. RISK is a 36-month prospective inception cohort study that has collected data from over 1000 pediatric Crohn’s disease patients from 28 clinics across the United States and Canada. Clinical, demographic and genetic data is available. In particular, we leveraged the recorded disease severity (wPCDAI scores at enrollment) and genotyping data (Infinium Global Screening Array) on 405 Crohn’s disease RISK patients who had both data types recorded. The weighted Pediatric Crohn’s Disease Activity Index (wPCDAI) consists of multiple subcomponents such as abdominal pain, stool frequency and extraintestinal manifestations to score disease severity in pediatric IBD. We performed imputation of the RISK genetic data to obtain genotypes on 12.1 million variants. Finally, we computed pPRS values across all 31 pathways as well as cPRS for all patients.

We first tested for correlation between cPRS and disease severity (Figure 6). We assigned patients into four severity bins as defined by Turner et al. [23] based on their wPCDAI at enrollment (remission: wPCDAI < 12.5, mild disease: 12.5-40, moderate: 40-60, severe > 60). Conventional PRS values exhibited a significant increasing trend as disease severity increased (Jonckheere-Terpstra trend test p = 0.034). Subcomponents used in the calculation of wPCDAI also showed an association with cPRS: limitation of activity was more pronounced with increasing cPRS values (Jonckheere-Terpstra trend test p = 0.006) (Figure 6). These statistically significant connections between cPRS and disease severity in the RISK cohort highlight the ability of polygenic risk scores to identify biologically relevant genetic risk burden within all-patient cohorts.

**Figure 6.**
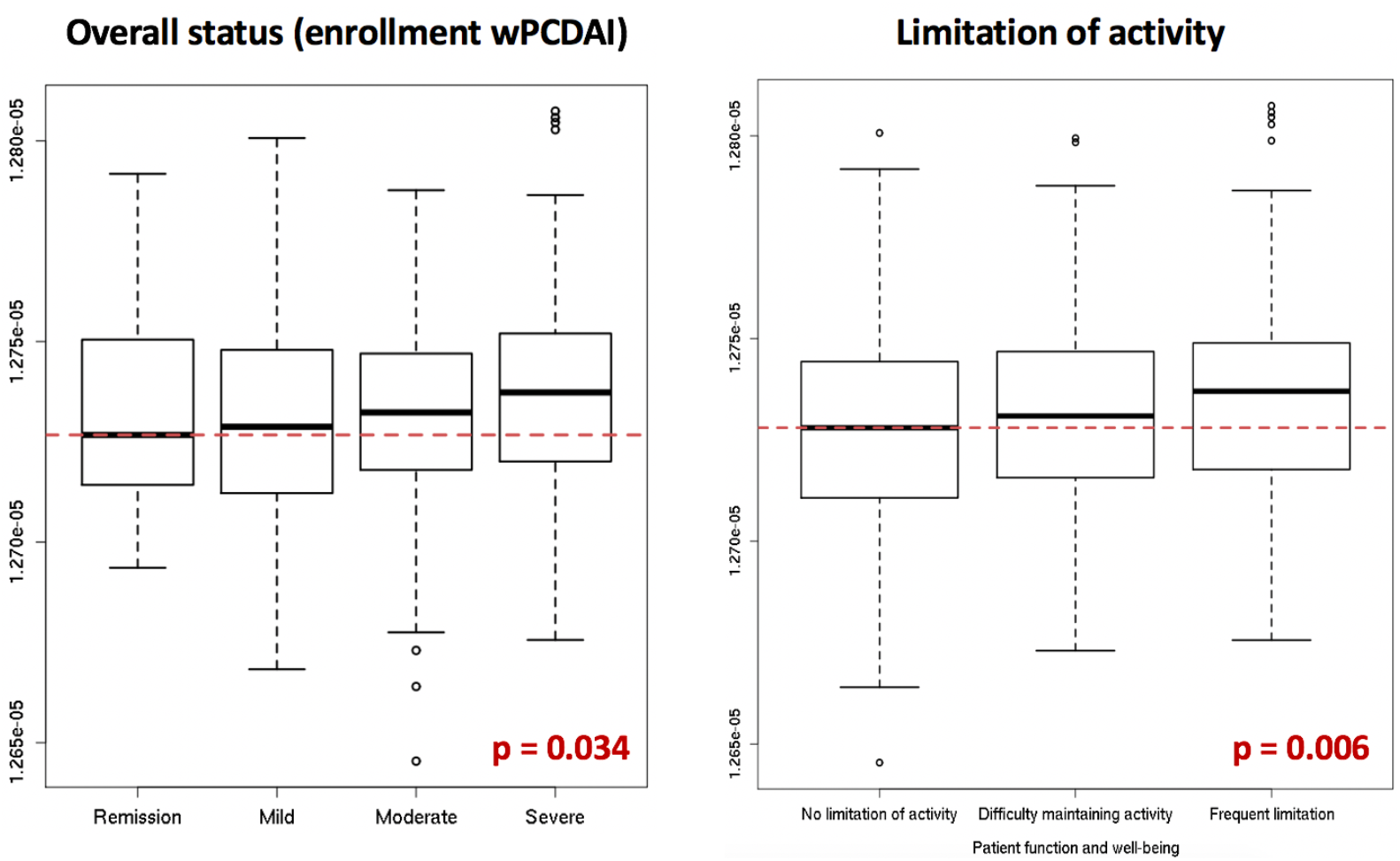
IBD cPRS scores for RISK patients across increasing levels of disease severity at enrollment. Increasing categories of disease severity correlated with higher cPRS values (wPCDAI metric: Jonckheere-Terpstra trend test p = 0. 034; limitation of activity: Jonckheere-Terpstra trend test p = 0.006).

Next, we computed pPRS values for all 405 patients with available genotyping data and wPCDAI scores. Elastic net logistic regression was used to identify the pathways most associated with wPCDAI (Figure 7). Eight pathways were identified as significant features by the elastic net model, and all coefficients were positive. This suggested that IBD driven by higher genetic risk burden within these pathways correlated with a more severe disease manifestation. Hierarchical clustering of the RISK samples based on pPRS values also correlated with disease severity: the first level of clustering identified a group of patients enriched for wPCDAI values > 30 (hypergeometric test p-value = 0.019) (Figure 7). These results showcased that pPRS can identify statistically significant connections between individual pathways and disease severity, and also identified patient subpopulations enriched for more severe manifestations.

**Figure 7.**
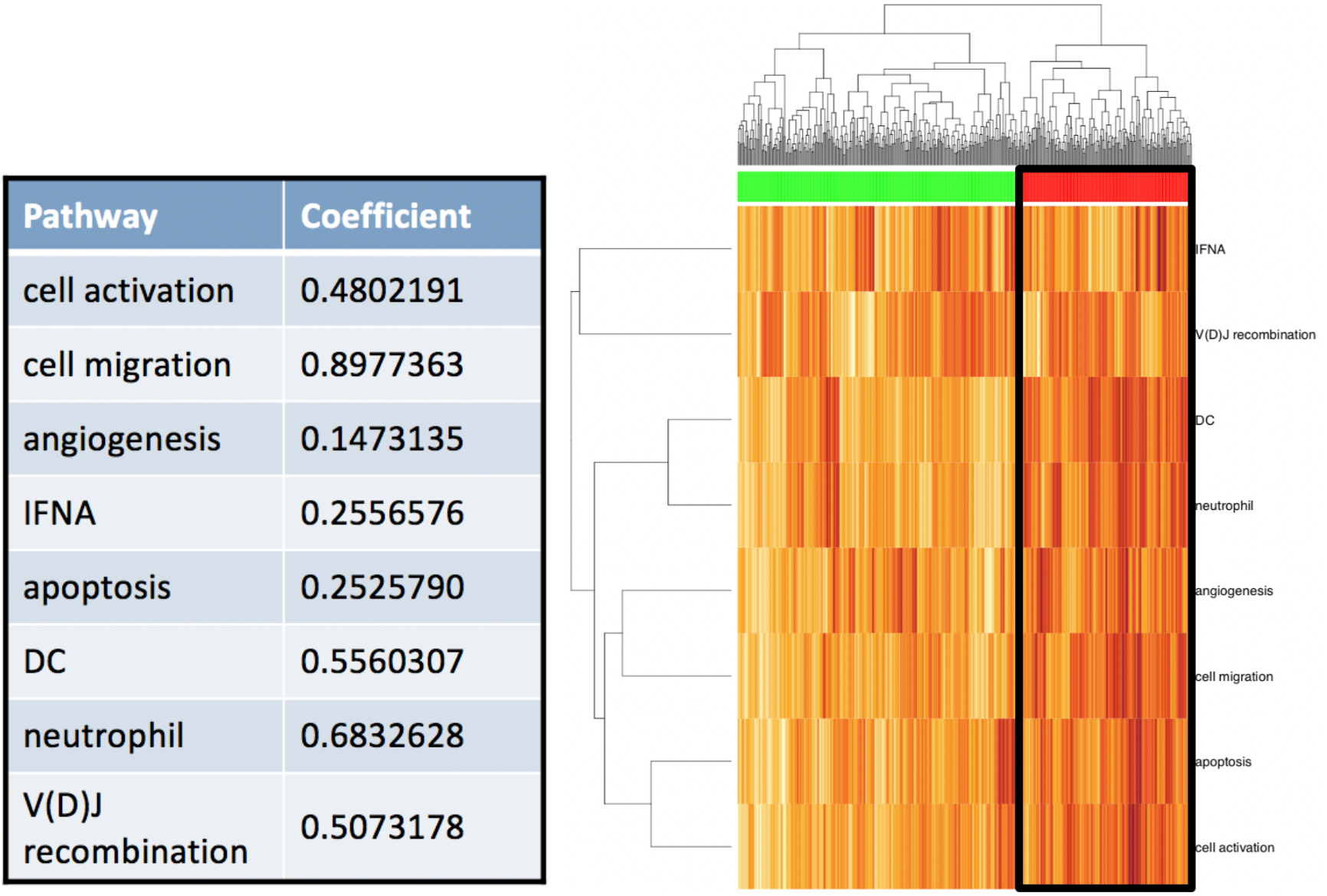
IBD severity pathways prioritized by elastic net logistic regression. Eight pathways were prioritized by the regularized regression model, with all pathways showing a positive association between genetic risk and disease severity. Hierarchical clustering based on these eight pathways identified a RISK patient cohort enriched for wPCDAI values > 30 (red cluster: hypergeometric test p-value = 0.019).

### Pathway analysis of IBD complications and anti-TNF treatment response

Two additional characteristics of an IBD disease course are the presence of complications (intestinal strictures, abscesses or fistulas) and response to anti-TNF treatment. To evaluate the connection between pPRS values and these disease characteristics we leveraged IBD patient data from the personalized anti-TNF therapy in Crohn’s disease study (PANTS) [17]. PANTS is a prospective observational UK-wide study that enrolled anti-TNF naïve Crohn’s disease patients. After anti-TNF treatment, patients were evaluated for 12 months or until drug withdrawal, and clinical as well as genetic data was collected at baseline. Patients were then evaluated for primary non-response.

We first tested for an association between IBD complications and pPRS values. We analyzed data from a total of 1304 PANTS patients who had both genetic and clinical data recorded: 779 patients in the B1 category (non-stricturing, non-penetrating disease), and 525 patients in the B2 (stricturing disease) or B3 category (penetrating disease). An elastic net logistic regression was run to model B1 versus B2 or B3 disease course, and 9 pathways as well as smoking history were identified by the regularized model as associated with IBD complications (Table 2). All pPRS coefficients were positive, indicating that higher genetic risk burden over these pathways correlated to higher likelihood of disease complications. Smoking (encoded as never, past, or current smoker) was also found to be positively correlated to incidence of Crohn’s disease complications, an observation that is supported by multiple studies [24,25,26,27,28] and serves as a positive control in our model.

**Table 2.**
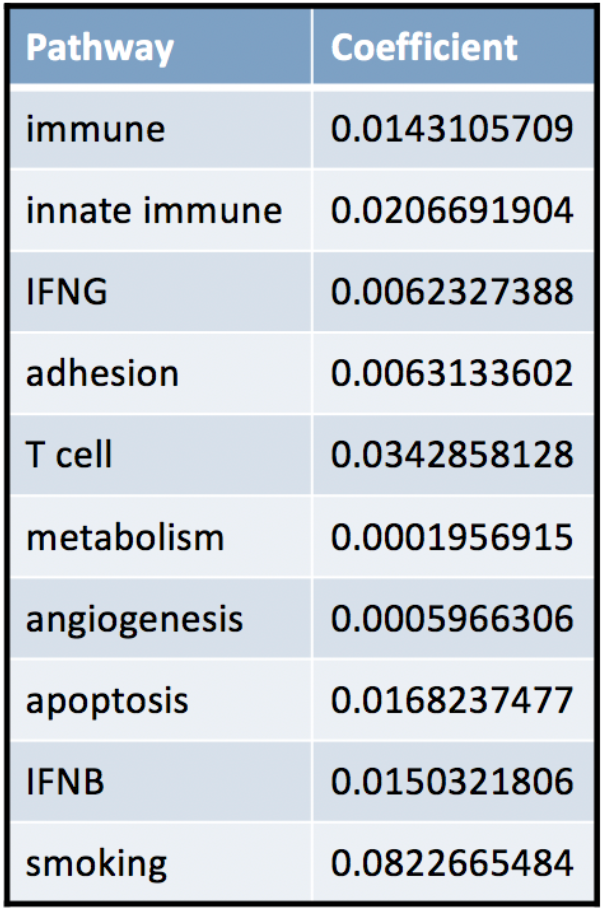
Pathways associated with IBD complications (B1 vs. B2 or B3 disease classification) identified by elastic net logistic regression. Nine pPRS features as well as smoking were prioritized by an elastic net regularized regression model of IBD complications. All features were positively associated with an increased likelihood of stricturing or penetrating IBD phenotype.

Finally, we analyzed the relationship between pPRS values and response to anti-TNF treatment in IBD. Within our patient cohort we observed 227 responders and 606 primary non-responders. Performing elastic net logistic regression on responder versus non-responder status identified 8 pathways associated with response, as well as smoking history. The regression model reported positive as well as negative coefficients, suggesting that our pPRS approach identified disease-relevant biological mechanisms that correlate positively as well as negatively with anti-TNF treatment response. The connection between the complement activation pathway and IBD susceptibility as well as anti-TNF treatment response across multiple diseases has been particularly well documented [29,30] and was also identified in our analysis. We also observed a negative association between smoking status and treatment response, which is concordant with previous studies across multiple autoimmune diseases [31,32,33,34,35].

We also applied our pPRS methodology to identify clusters of PANTS IBD patients that were enriched or depleted for responders to anti-TNF treatment (Figure 8). Starting from the overall cohort composition (73% responders, 27% non-responders) we observed that stratifying the PANTS cohort based on the extremes of cPRS values (top or bottom 20%) did not lead to strong changes in response fractions. Stratifying the PANTS cohort by smoking status shows that never smokers contained a higher fraction of responders (76%) and smokers (past or current) contained a higher fraction of non-responders (31%) compared to the PANTS cohort average. Adding information on pPRS values further improved the stratification. Combining a history of never smoking with high complement activation pPRS values (top 20%) led to a patient subset with a higher responder fraction compared to the overall cohort (81% versus 73%). On the other hand, combining a history of smoking (past or current) with low complement activation pPRS values (bottom 20%) led to a patient subset with a higher non-responder fraction compared to the overall cohort (40% versus 27%). The complement activation pathway has been connected to IBD susceptibility [29] as well as response to anti-TNF treatment [30], and in line with these previous results it is also a hit in our susceptibility analysis (the complement activation pathway was identified in our analysis of IBD-relevant genes) as well as in our treatment response analysis: this pathway was identified by the elastic net logistic regression model (Table 3) and helped enrich for responders or non-responders in our patient stratification analysis (Figure 8). Other pathways identified by the elastic net model were also leveraged for patient stratification (for example the hemopoiesis pathway results in Figure 8), although stratifying by multiple pathways simultaneously could lead to a drop in sample sizes.

**Table 3.** Pathways associated with anti-TNF treatment response identified by elastic net logistic regression. Eight pPRS features as well as smoking were prioritized by an elastic net regularized regression model of anti-TNF treatment response in PANTS patients. Pathway PRS features had a mix of positive and negative regression coefficients, indicating different areas of disease biology and their individual relationship to treatment response. A history of smoking correlated negatively with treatment response.

| Pathway | Coefficient |
| --- | --- |
| IFNG | -0.019350993 |
| hemopoiesis | 0.074602649 |
| angiogenesis | -0.016183419 |
| inactivation of innate immune signaling | 0.001403924 |
| calcium ion transport in T cells | -0.080374711 |
| phagocytic superoxide | 0.015554464 |
| complement activation | 0.052155737 |
| alkaline phosphatase | -0.004337945 |
| smoking | -0.128544286 |

**Figure 8.**
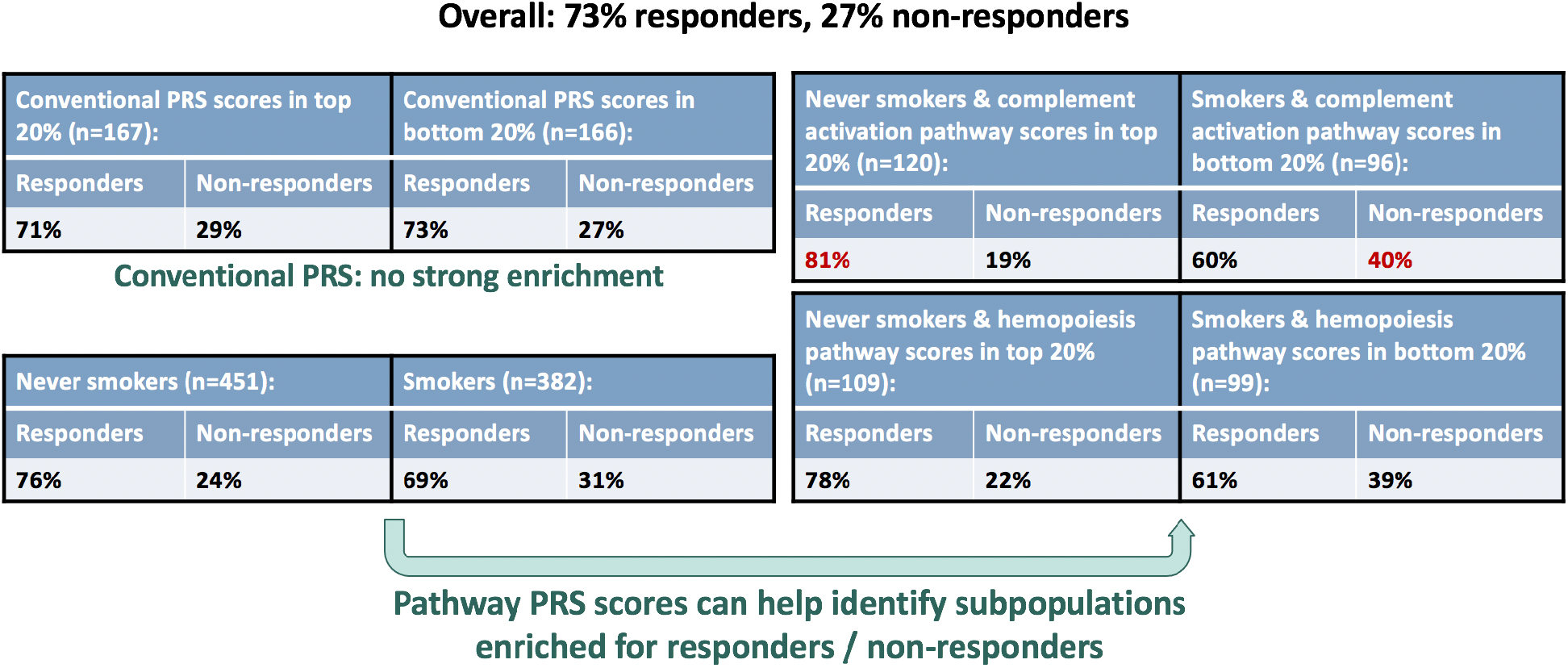
Identifying patient subpopulations enriched for anti-TNF responders or non-responders based on pathway PRS scores. Fractions of responders and non-responders and sample sizes are listed for the full PANTS cohort as well as for patient clusters defined based on smoking history and pPRS scores for hemopoiesis and complement activation.

## Discussion

We propose new methodology for pathway-centered polygenic risk score models that leverages transcriptomic, epigenetic and functional annotations in the form of Open Targets V2G scores to connect PRS variants to relevant genes, and defines pathways starting from a carefully prioritized list of genes with robust GWAS and Mendelian evidence for disease involvement. We have extensively assessed the potential for this pPRS model to identify relevant IBD biology (susceptibility, severity, complications) and to aid in patient stratification. We also evaluated whether this pPRS model informed on biology related to anti-TNF treatment response by pointing to disease-relevant pathways that are associated with altered response levels. Present findings suggest that pathway-based risk scores are associated with disease behavior as well as treatment response. These risk scores also have some power to stratify patients although this stratification performs best at the extremes of the risk score distribution (high or low-scoring patients).

Our analysis of three complementary IBD datasets allowed us to study multiple aspects of disease biology. The analysis of IBD cases and controls collected in the UK Biobank confirmed that most of our pathways were strongly and positively associated with disease susceptibility. While this result was expected given the construction of our model it showcased that a pPRS approach identified biologically relevant signals and accurately reported on effect directions. Our UK Biobank analysis also highlighted that sample clustering could be performed based on pathways prioritized by an elastic net model, and that this clustering can significantly enrich for IBD cases or controls.

Our analysis of early-onset IBD cases in the RISK cohort connected eight pathways to disease severity, with higher genetic risk burden over these pathways correlating with more severe manifestations. Hierarchical patient clustering based on these eight pathways also identified groups enriched for severe cases (based on the wPCDAI metric). Disease complications (stricturing or penetrating IBD phenotypes) were also connected to individual pathways based on our analysis of the PANTS cohort, and smoking was found to correlate with the presence of IBD complications. Finally, multiple pathways were positively or negatively associated with anti-TNF treatment response, suggesting different areas of disease biology that may influence drug response. Using pPRS as a patient stratification metric we were able to create patient cohorts enriched for responders or non-responders based solely on the genetic data encoded by the polygenic risk score. In addition, pPRS-based stratification led to higher fractions of responders or non-responders in the corresponding PANTS subgroups than a stratification based on cPRS values.

Both conventional and pathway PRS methodologies rely heavily on common variant genetic evidence provided by large-scale GWAS studies. An integration of common variant evidence with rare variant burden test results has the potential to capture additional genetic contribution to disease and further improve our statistical power to connect biological pathways to genetically influenced traits. As larger cohorts facilitate an improved identification of genetic associations at the rare and common variant level, pPRS methods will be useful tools for both therapeutic-focused pathway prioritization as well as patient stratification.

## Data Availability

All data produced in the present work are contained in the manuscript.

## Acknowledgements

The results published here are in whole or part based on data from the RISK pediatric CD cohort study obtained from the IBD Plexus program of the Crohn’s & Colitis Foundation (funding was provided by AbbVie for the IBD Plexus Program).

This research has been conducted using the UK Biobank Resource under Application Number 26041.

## Supplemental information

**Figure S1.**
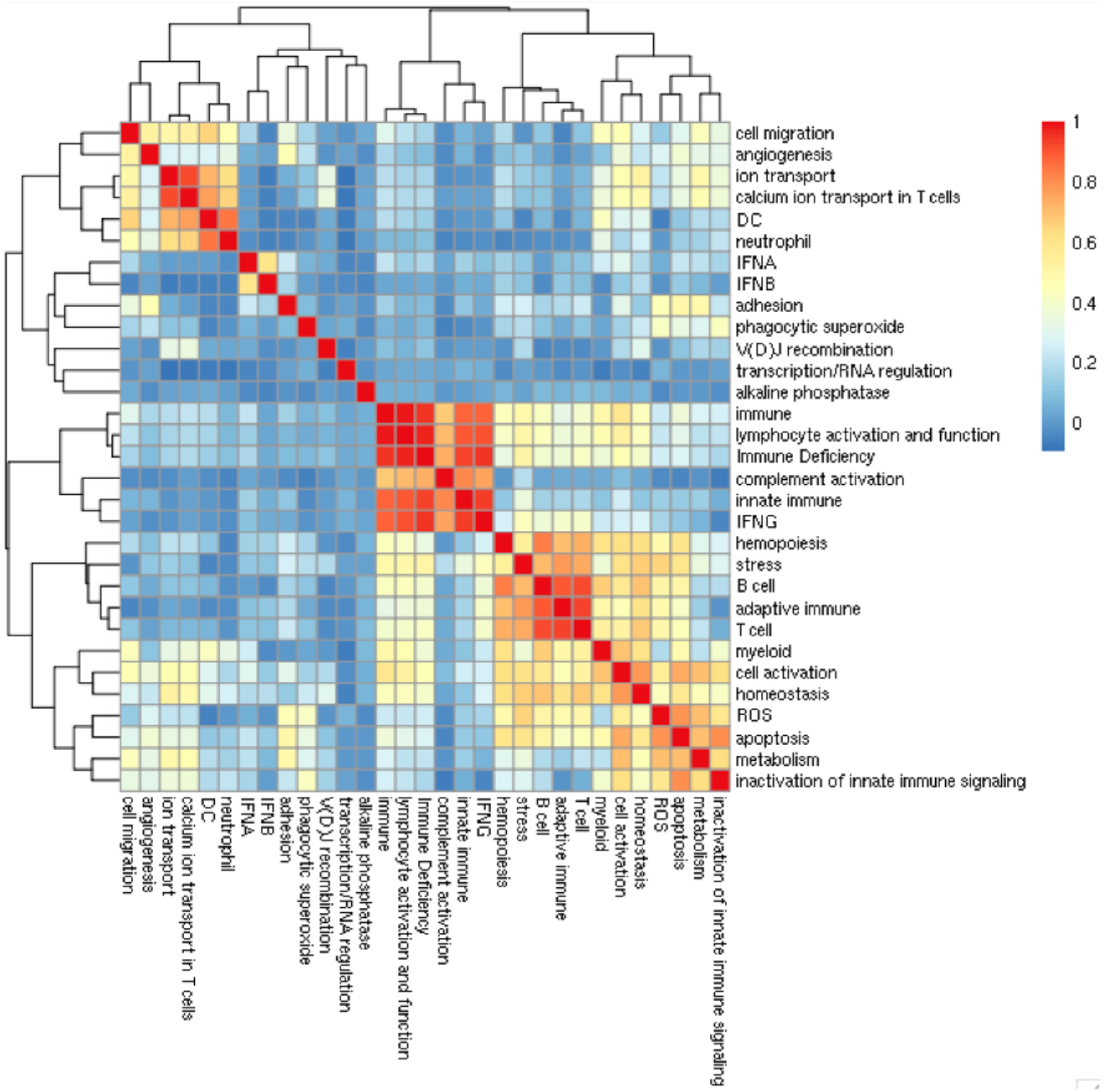
Correlation structure of pPRS scores on UK Biobank samples. The observed absolute values of Pearson correlation coefficients reflect shared genes present in multiple pathways.

## References

1. Liu JZ, Anderson CA. Genetic studies of Crohn’s disease: past, present and future. Best Pract Res Clin Gastroenterol. 2014 Jun;28(3):373–86. doi: 10.1016/j.bpg.2014.04.009. Epub 2014 May 6. PMID: 24913378; PMCID: PMC4075408.

2. Moller FT, Andersen V, Wohlfahrt J, Jess T. Familial risk of inflammatory bowel disease: a population-based cohort study 1977-2011. Am J Gastroenterol. 2015 Apr;110(4):564–71. doi: 10.1038/ajg.2015.50. Epub 2015 Mar 24. PMID: 25803400.

3. Podolsky D.K. Inflammatory Bowel Disease. N Engl J Med 2002; 347:417–429

4. Uhlig H.H. Monogenic diseases associated with intestinal inflammation: implications for the understanding of inflammatory bowel disease. Gut 2013; 62:1795–1805.

5. Jodie Ouahed, MD MMSc, Elizabeth Spencer, M., Daniel Kotlarz, MD PhD, Dror S Shouval, MD MMSc, Matthew Kowalik, M., Kaiyue Peng, M., Michael Field, M., Leslie Grushkin-Lerner, PhD, Sung-Yun Pai, MD, Athos Bousvaros, MD, MPH, Judy Cho, M., Carmen Argmann, PhD, Eric Schadt, PhD, Dermot P B McgovernMD, PhD, Michal MokryMD, PhD, Edward NieuwenhuisMD, PhD, Hans CleversMD, PhD, Fiona Powrie, DPhil, Holm Uhlig, MD, D Phil, Christoph KleinMD, PhD, Aleixo Muise, MD PhD, Marla Dubinsky, M., Scott B Snapper, MD PhD, Very Early Onset Inflammatory Bowel Disease: A Clinical Approach With a Focus on the Role of Genetics and Underlying Immune Deficiencies, Inflammatory Bowel Diseases, Volume 26, Issue 6, June 2020, Pages 820–842.

6. Holm H. Uhlig, Tobias Schwerd, Sibylle Koletzko, Neil Shah, Jochen Kammermeier, Abdul Elkadri, Jodie Ouahed, David C. Wilson, Simon P. Travis, Dan Turner, Christoph Klein, Scott B. Snapper, Aleixo M. Muise. The Diagnostic Approach to Monogenic Very Early Onset Inflammatory Bowel Disease. Gastroenterology, Volume 147, Issue 5, 2014, Pages 990–1007.e3.

7. de Lange KM, Moutsianas L, Lee JC, Lamb CA, Luo Y, Kennedy NA, Jostins L, Rice DL, Gutierrez-Achury J, Ji SG, Heap G, Nimmo ER, Edwards C, Henderson P, Mowat C, Sanderson J, Satsangi J, Simmons A, Wilson DC, Tremelling M, Hart A, Mathew CG, Newman WG, Parkes M, Lees CW, Uhlig H, Hawkey C, Prescott NJ, Ahmad T, Mansfield JC, Anderson CA, Barrett JC. Genome-wide association study implicates immune activation of multiple integrin genes in inflammatory bowel disease. Nat Genet. 2017 Feb;49(2):256–261.

8. Huang, H., Fang, M., Jostins, L. et al. Fine-mapping inflammatory bowel disease loci to single-variant resolution. Nature 547, 173–178 (2017).

9. Giambartolomei C, Vukcevic D, Schadt EE, Franke L, Hingorani AD, Wallace C, et al. (2014) Bayesian Test for Colocalisation between Pairs of Genetic Association Studies Using Summary Statistics. PLoS Genet 10(5): e1004383. https://doi.org/10.1371/journal.pgen.1004383

10. Khera, A.V., Chaffin, M., Aragam, K.G. et al. Genome-wide polygenic scores for common diseases identify individuals with risk equivalent to monogenic mutations. Nat Genet 50, 1219–1224 (2018). https://doi.org/10.1038/s41588-018-0183-z

11. Ahmad S, Bannister C, van der Lee SJ, Vojinovic D, Adams HHH, Ramirez A, Escott-Price V, Sims R, Baker E, Williams J, Holmans P, Vernooij MW, Ikram MA, Amin N, van Duijn CM. Disentangling the biological pathways involved in early features of Alzheimer’s disease in the Rotterdam Study. Alzheimers Dement. 2018 Jul;14(7):848–857. doi: 10.1016/j.jalz.2018.01.005. Epub 2018 Mar 1. PMID: 29494809.

12. Darst BF, Koscik RL, Racine AM, Oh JM, Krause RA, Carlsson CM, Zetterberg H, Blennow K, Christian BT, Bendlin BB, Okonkwo OC, Hogan KJ, Hermann BP, Sager MA, Asthana S, Johnson SC, Engelman CD. Pathway-Specific Polygenic Risk Scores as Predictors of Amyloid-β Deposition and Cognitive Function in a Sample at Increased Risk for Alzheimer’s Disease. J Alzheimers Dis. 2017;55(2):473–484. doi: 10.3233/JAD-160195. PMID: 27662287; PMCID: PMC5123972.

13. Sandling JK, Pucholt P, Hultin Rosenberg L, et al. Molecular pathways in patients with systemic lupus erythematosus revealed by gene-centred DNA sequencing. Annals of the Rheumatic Diseases 2021;80:109–117.

14. Maya Ghoussaini, Edward Mountjoy, Miguel Carmona, Gareth Peat, Ellen M Schmidt, Andrew Hercules, Luca Fumis, Alfredo Miranda, Denise Carvalho-Silva, Annalisa Buniello, Tony Burdett, James Hayhurst, Jarrod Baker, Javier Ferrer, Asier Gonzalez-Uriarte, Simon Jupp, Mohd Anisul Karim, Gautier Koscielny, Sandra Machlitt-Northen, Cinzia Malangone, Zoe May Pendlington, Paola Roncaglia, Daniel Suveges, Daniel Wright, Olga Vrousgou, Eliseo Papa, Helen Parkinson, Jacqueline A L MacArthur, John A Todd, Jeffrey C Barrett, Jeremy Schwartzentruber, David G Hulcoop, David Ochoa, Ellen M McDonagh, Ian Dunham, Open Targets Genetics: systematic identification of trait-associated genes using large-scale genetics and functional genomics, Nucleic Acids Research, Volume 49, Issue D1, 8 January 2021, Pages D1311–D1320.

15. Bycroft, C., Freeman, C., Petkova, D. et al. The UK Biobank resource with deep phenotyping and genomic data. Nature 562, 203–209 (2018). https://doi.org/10.1038/s41586-018-0579-z

16. Walters TD, Kim MO, Denson LA, Griffiths AM, Dubinsky M, Markowitz J, Baldassano R, Crandall W, Rosh J, Pfefferkorn M, Otley A, Heyman MB, LeLeiko N, Baker S, Guthery SL, Evans J, Ziring D, Kellermayer R, Stephens M, Mack D, Oliva-Hemker M, Patel AS, Kirschner B, Moulton D, Cohen S, Kim S, Liu C, Essers J, Kugathasan S, Hyams JS; PRO-KIIDS Research Group. Increased effectiveness of early therapy with anti-tumor necrosis factor-α vs an immunomodulator in children with Crohn’s disease. Gastroenterology. 2014 Feb;146(2):383–91. doi: 10.1053/j.gastro.2013.10.027. Epub 2013 Oct 23. PMID: 24162032.

17. Kennedy NA, Heap GA, Green HD, Hamilton B, Bewshea C, Walker GJ, Thomas A, Nice R, Perry MH, Bouri S, Chanchlani N, Heerasing NM, Hendy P, Lin S, Gaya DR, Cummings JRF, Selinger CP, Lees CW, Hart AL, Parkes M, Sebastian S, Mansfield JC, Irving PM, Lindsay J, Russell RK, McDonald TJ, McGovern D, Goodhand JR, Ahmad T; UK Inflammatory Bowel Disease Pharmacogenetics Study Group. Predictors of anti-TNF treatment failure in anti-TNF-naive patients with active luminal Crohn’s disease: a prospective, multicentre, cohort study. Lancet Gastroenterol Hepatol. 2019 May;4(5):341–353. doi: 10.1016/S2468-1253(19)30012-3. Epub 2019 Feb 27. PMID: 30824404.

18. Yang J, Ferreira T, Morris AP, Medland SE; Genetic Investigation of ANthropometric Traits (GIANT) Consortium; DIAbetes Genetics Replication and Meta-analysis (DIAGRAM) Consortium, Madden PA, Heath AC, Martin NG, Montgomery GW, Weedon MN, Loos RJ, Frayling TM, McCarthy MI, Hirschhorn JN, Goddard ME, Visscher PM. Conditional and joint multiple-SNP analysis of GWAS summary statistics identifies additional variants influencing complex traits. Nat Genet. 2012 Mar 18;44(4):369–75, S1-3. doi: 10.1038/ng.2213. PMID: 22426310; PMCID: PMC3593158.

19. GTEx Consortium. The GTEx Consortium atlas of genetic regulatory effects across human tissues. Science. 2020 Sep 11;369(6509):1318–1330. doi: 10.1126/science.aaz1776. PMID: 32913098; PMCID: PMC7737656.

20. Szklarczyk D, Gable AL, Lyon D, Junge A, Wyder S, Huerta-Cepas J, Simonovic M, Doncheva NT, Morris JH, Bork P, Jensen LJ, Mering CV. STRING v11: protein-protein association networks with increased coverage, supporting functional discovery in genome-wide experimental datasets. Nucleic Acids Res. 2019 Jan 8;47(D1):D607–D613. doi: 10.1093/nar/gky1131. PMID: 30476243; PMCID: PMC6323986.

21. Ashburner M, Ball CA, Blake JA, Botstein D, Butler H, Cherry JM, Davis AP, Dolinski K, Dwight SS, Eppig JT, Harris MA, Hill DP, Issel-Tarver L, Kasarskis A, Lewis S, Matese JC, Richardson JE, Ringwald M, Rubin GM, Sherlock G. Gene ontology: tool for the unification of biology. The Gene Ontology Consortium. Nat Genet. 2000 May;25(1):25–9. doi: 10.1038/75556. PMID: 10802651; PMCID: PMC3037419.

22. Gene Ontology Consortium. The Gene Ontology resource: enriching a GOld mine. Nucleic Acids Res. 2021 Jan 8;49(D1):D325–D334. doi: 10.1093/nar/gkaa1113. PMID: 33290552; PMCID: PMC7779012.

23. Dan Turner, MD PhD, Anne M. Griffiths, MD, Thomas D. Walters, MD, Tong Seah, James Markowitz, M., Marian Pfefferkorn, M., David Keljo, M., Jacob Waxman, Anthony Otley, MD MSc, Neal S. LeLeiko, MD PhD, David Mack, M., Jeffrey Hyams, M., Arie Levine, MD, Mathematical Weighting of the Pediatric Crohn’s Disease Activity Index (PCDAI) and Comparison with Its Other Short Versions, Inflammatory Bowel Diseases, Volume 18, Issue 1, 1 January 2012, Pages 55–62.

24. Gustavsson A, Magnuson A, Blomberg B, Andersson M, Halfvarson J, Tysk C. Smoking is a risk factor for recurrence of intestinal stricture after endoscopic dilation in Crohn’s disease. Aliment Pharmacol Ther. 2013 Feb;37(4):430–7. doi: 10.1111/apt.12176. Epub 2012 Dec 3. PMID: 23205619.

25. Thomas GA, Rhodes J, Green JT, Richardson C. Role of smoking in inflammatory bowel disease: implications for therapy. Postgrad Med J. 2000 May;76(895):273–9. doi: 10.1136/pmj.76.895.273. PMID: 10775279; PMCID: PMC1741576.

26. Gareth C. Parkes, Kevin Whelan, James O. Lindsay, Smoking in inflammatory bowel disease: Impact on disease course and insights into the aetiology of its effect, Journal of Crohn’s and Colitis, Volume 8, Issue 8, 1 August 2014, Pages 717–725.

27. Lakatos PL, Vegh Z, Lovasz BD, David G, Pandur T, Erdelyi Z, Szita I, Mester G, Balogh M, Szipocs I, Molnar C, Komaromi E, Golovics PA, Mandel M, Horvath A, Szathmari M, Kiss LS, Lakatos L. Is current smoking still an important environmental factor in inflammatory bowel diseasesã Results from a population-based incident cohort. Inflamm Bowel Dis. 2013 Apr;19(5):1010–7. doi: 10.1097/MIB.0b013e3182802b3e. PMID: 23399739.

28. Lawrance IC, Murray K, Batman B, Gearry RB, Grafton R, Krishnaprasad K, Andrews JM, Prosser R, Bampton PA, Cooke SE, Mahy G, Radford-Smith G, Croft A, Hanigan K. Crohn’s disease and smoking: is it ever too late to quitã J Crohns Colitis. 2013 Dec;7(12):e665–71. doi: 10.1016/j.crohns.2013.05.007. Epub 2013 Jun 20. PMID: 23790611.

29. Nissilä E, Korpela K, Lokki AI, Paakkanen R, Jokiranta S, de Vos WM, Lokki ML, Kolho KL, Meri S. C4B gene influences intestinal microbiota through complement activation in patients with paediatric-onset inflammatory bowel disease. Clin Exp Immunol. 2017 Dec;190(3):394–405. doi: 10.1111/cei.13040. Epub 2017 Sep 25. PMID: 28832994; PMCID: PMC5680072.

30. Sandahl TD, Kelsen J, Dige A, Dahlerup JF, Agnholt J, Hvas CL, Thiel S. The lectin pathway of the complement system is downregulated in Crohn’s disease patients who respond to anti-TNF-α therapy. J Crohns Colitis. 2014 Jun;8(6):521–8. doi: 10.1016/j.crohns.2013.11.007. Epub 2013 Nov 28. PMID: 24291022.

31. Söderlin MK, Petersson IF, Geborek P. The effect of smoking on response and drug survival in rheumatoid arthritis patients treated with their first anti-TNF drug. Scand J Rheumatol. 2012 Feb;41(1):1–9. doi: 10.3109/03009742.2011.599073. Epub 2011 Nov 28. PMID: 22118371.

32. Bente Glintborg, Pil Højgaard, Merete Lund Hetland, Niels Steen Krogh, Gina Kollerup, Jørgen Jensen, Stavros Chrysidis, Inger Marie Jensen Hansen, Mette Holland-Fischer, Torben Højland Hansen, Christine Nilsson, Jakob Espesen, Henrik Nordin, Anne Gitte Rasmussen Loft, Randi Pelck, Tove Lorenzen, Sussi Flejsborg Oeftiger, Barbara Unger, Frank Jaeger, Peter Mosborg Petersen, Claus Rasmussen, Lene Dreyer. Impact of tobacco smoking on response to tumour necrosis factor-alpha inhibitor treatment in patients with ankylosing spondylitis: results from the Danish nationwide DANBIO registry, Rheumatology, Volume 55, Issue 4, April 2016, Pages 659–668.

33. Abhishek A, Butt S, Gadsby K, Zhang W, Deighton CM. Anti-TNF-alpha agents are less effective for the treatment of rheumatoid arthritis in current smokers. J Clin Rheumatol. 2010 Jan;16(1):15–8. doi: 10.1097/RHU.0b013e3181ca4a2a. PMID: 20051749.

34. Hyrich KL, Watson KD, Silman AJ, Symmons DP; British Society for Rheumatology Biologics Register. Predictors of response to anti-TNF-alpha therapy among patients with rheumatoid arthritis: results from the British Society for Rheumatology Biologics Register. Rheumatology (Oxford). 2006 Dec;45(12):1558–65. doi: 10.1093/rheumatology/kel149. Epub 2006 May 16. PMID: 16705046.

35. Mattey DL, Brownfield A, Dawes PT. Relationship between pack-year history of smoking and response to tumor necrosis factor antagonists in patients with rheumatoid arthritis. J Rheumatol. 2009 Jun;36(6):1180–7. doi: 10.3899/jrheum.081096. Epub 2009 May 15. PMID: 19447930.

